# Seroprevalence of SARS-CoV-2, symptom profiles and seroneutralization during the first COVID-19 wave in a suburban area, France

**DOI:** 10.1101/2021.02.10.21250862

**Authors:** Anne Gégout petit, Hélène Jeulin, Karine Legrand, Agathe Bochnakian, Pierre Vallois, Evelyne Schvoerer, Francis Guillemin

**Affiliations:** Université de Lorraine, CNRS, Inria, IECL, F-54000 Nancy, France; Université de Lorraine, CNRS, LCPME, F-54000 Nancy, France; Laboratoire de Virologie, CHRU de Nancy Brabois, F-54500 Vandoeuvre-lès-Nancy, France; CHRU-Nancy, INSERM, Université de Lorraine, CIC Epidémiologie clinique, F-54000 Nancy, France

## Abstract

**Background:** The World Health Organisation recommends monitoring the circulation of severe acute respiratory syndrome coronavirus 2 (SARS-CoV-2). We aimed to estimate anti–SARS-CoV-2 total immunoglobulin (IgT) antibody seroprevalence and describe symptom profiles and *in vitro* seroneutralization in Nancy, France, in spring 2020.

**Methods:** Individuals were randomly sampled from electoral lists and invited with household members over 5 years old to be tested for anti–SARS-CoV-2 (IgT, i.e. IgA/IgG/IgM) antibodies by ELISA (Bio-rad). Serum samples were classified according to seroneutralization activity >50% (NT50) on Vero CCL-81 cells. Age- and sex-adjusted seroprevalence was estimated. Subgroups were compared by chi-square or Fisher exact test and logistic regression.

**Results:** Among 2006 individuals, 43 were SARS-CoV-2–positive; the raw seroprevalence was 2.1% (95% confidence interval 1.5 to 2.9), with adjusted metropolitan and national standardized seroprevalence 2.5% (1.8 to 3.3) and 2.3% (1.7 to 3.1). Seroprevalence was highest for 20-to 34-year-old participants (4.7% [2.3 to 8.4]), within than out of socially deprived area (2.5% vs 1%, *P=*0.02) and with than without intra-family infection (p<10^−6^). Moreover, 25% (23 to 27) of participants presented at least one COVID-19 symptom associated with SARS-CoV-2 positivity (p<10^−13^), with anosmia or ageusia highly discriminant (odds ratio 27.8 [13.9 to 54.5]), associated with dyspnea and fever. Among the SARS-CoV-2-positives, 16.3% (6.8 to 30.7) were asymptomatic. For 31 of these individuals, positive seroneutralization was demonstrated *in vitro*.

**Conclusions:** In this population of very low anti-SARS-CoV-2 antibody seroprevalence, a beneficial effect of the lockdown can be assumed, with frequent SARS-CoV-2 seroneutralization among IgT-positive patients.

**Key Messages:**

- Total immunoglobulin antibody (IgT) measurement is an accurate tool to monitor the circulation of severe acute respiratory syndrome coronavirus 2 (SARS-CoV-2) and a key biological feature to assume the spread of COVID-19 later after the appearance of symptoms.
- IgT seroprevalence was 2.1% in the Grand Nancy Metropolitan area, France; was highest for young adults; in socially deprived area, but this was not confirmed at the individual level; and was associated with high intra-family viral transmission.
- About two thirds of IgT-positive individuals exhibited SARS-CoV-2–positive seroneutralization.

**Trial registration:** NCT04448769

## BACKGROUND

The World Health Organisation (WHO) (1) recommends a good observation of the circulation of the severe acute respiratory syndrome coronavirus 2 (SARS-CoV-2), including local seroprevalence surveys, to adapt the public health response to COVID-19 (2). Indeed, population containment, sanitary procedures and planning must be defined in terms of a quantified health concern. To estimate the proportion of individuals who were or are infected by the virus, serology assays for detecting anti-SARS-CoV-2 antibodies are useful in all individuals with mild (or no) clinical signs, with or without a RT-PCR test.

Between January and July 2020, 13 general-population serology surveys of SARS-CoV-2 were reported in Europe, 10 in the United States, 4 in Brazil, 1 in Pakistan and 1 in Japan (*personal communication*). Most (n=20) estimated the seroprevalence between 0 and 5%; half under 2.5%. Six studies conducted in regions highly affected by the epidemic estimated the anti– SARS-CoV-2 antibody seroprevalence at more than 15% (2–7). Few studies investigated the relation between seroprevalence and social precariousness, despite some evidence that health inequalities are reflected in the pandemic (8,9).

Serology assays usually detect antibodies against the viral spike “S” and nucleocapsid “N” protein, both being highly antigenic and widely expressed during SARS-CoV-2 infection (10). After primary infection, immunoglobulin G (IgG) levels increase continuously, peaking at about 6 weeks after infection and often remaining high for 6 months. The neutralizing activity usually peaks after 4 weeks and then can slowly decrease (11). At 4 months after a first positive anti–SARS-CoV-2 antibody result, 41% of infected patients become negative for anti-N antibodies, most still positive for anti-S antibodies (12). The viral infection requires the receptor-binding domain (RBD) of the S protein, which is the molecular determinant of viral attachment to the host cell receptor angiotensin-converting enzyme 2 (ACE-2) (13). Antibodies targeting the S protein neutralize the virus entering into the cell, and the IgM, IgA and IgG antibodies directed at the RBD of the S protein are highly neutralizing (14–16). Thus, well-standardized, reproducible antibody assays are crucial to establish correlates of risk and protection so that SARS-CoV-2 neutralization assays can be used for antibody monitoring in natural infection and vaccine trials (17).

The first COVID-19 cases were reported in France in January 2020 (18) and a strong SARS-CoV-2 emergence was observed in northeast France in March and April 2020, with numerous patients presenting at Nancy University hospital, France (19). To document the strength of SARS-CoV-2 circulation, biological samples from a random sample of the population were needed for serological testing (1). Our primary objective was to estimate the anti–SARS-CoV-2 total Ig (IgT) antibody seroprevalence in a random sample of the population of the Grand Nancy Metropolitan area. The secondary objectives were to estimate 1) the proportion of asymptomatic cases or symptom profiles, 2) the proportion of seropositive people according to level of social precariousness, and 3) the *in vitro* neutralization capacity of viral infectivity for the detected anti–SARS-CoV-2 antibodies.

## METHODS

The COVAL Nancy cross-sectional study was conducted between 26 June and 24 July 2020.

### Sampling

The target population consisted of all inhabitants of the Grand Nancy metropolitan area who were ≥ 5 years old on 1 June 2020. Adults randomly sampled from the electoral lists were invited to participate with all household members. To ensure representativeness, sampling was carried out by strata of homogeneous housing areas according to socio-economic criteria (IRIS habitat; INSEE Source(s): INSEE, Géographie à l’infra-communale [Official Geographic Code]), with each homogeneous housing area (IRIS) associated with the European deprivation index (EDI) (20). This continuous index consists of ecological variables best identified to reflect the individual experience of deprivation and is grouped by INSEE into classes by quintiles; 5 is the most deprived class.

From preliminary regional estimates with strong county disparities (21) and given serologic test sensitivity (100%) and specificity (99.5%), with 1% target precision and 95% confidence interval, we needed 1987 individuals to detect a 5% seroprevalence. Accordingly, the survey logistics were organized to account for estimated individual response rate, household members’ participation, appointment attendance and agreeing for blood sampling; we aimed to include 2000 individuals.

All invited individuals were informed of the objectives and the workflow of the study by using comprehensive messaging adapted to age. All individuals gave their signed consent.

Ethical approval was obtained (Comité de Protection des Personnes EST III, NANCY, France: ID RCB 2020-A01593-36) on 16 June 2020 and the French Commission for Individual Data Protection and Public Liberties (CNIL) on 19 June 2020.

During the inclusion visit to the Nancy University Hospital, each participant completed a self-reporting questionnaire adapted to age (adult, adolescent; child questionnaire completed by parents). The following data were collected:

- socio-demographic characteristics: age, sex, socio-professional category, education level;
- Evaluation of Deprivation and Inequalities in Health Examination Centres (EPICES) questionnaire (for adults) (22), a composite index commonly used to measure individual deprivation. A score is calculated on the basis of 11 weighted questions related to material and social deprivation, ranging from 0 to 100 (> 30 associated with social deprivation).
- health characteristics: body mass index, smoking status, influenza vaccination, health problems, pregnancy;
- potential contacts with a person with COVID-19: perception of infection with the virus, relatives infected;
- symptoms experienced since mid-February: fever, cough, runny nose, chest pain, anosmia or ageusia, sore throat, muscle pain, aches, fatigue, headaches, skin rashes, appetite loss, shortness of breath, diarrhoea, loss of balance, abdominal pain, nausea, and irritated eyes. According to the European Center for Disease Prevention and Control (23), at least one symptom among fever, cough, anosmia or ageusia, and shortness of breath indicates COVID-19.

### Serology

Blood samples were centrifuged to collect serum, which was stored at +4°C and then −20°C. Total anti-SARS-CoV-2 antibodies (IgA/IgG/IgM) were detected by using ELISA (Platelia SARS-CoV-2 Total Ab Assay, Bio-rad) on an Evolis Premium device (Bio-rad). IgM and IgG antibodies were detected by using an imunochromatographic test (Biosynex), with IgA by ELISA (Anti-SARS-CoV-2 Assay, Euroimmun) on SARS-CoV-2–seropositive samples and on samples from SARS-CoV-2–seronegative individuals living with SARS-CoV-2–positive individuals.

A person was classified as SARS-CoV-2–seropositive if at least two serology tests were positive (Figure 1).

**Figure 1.**
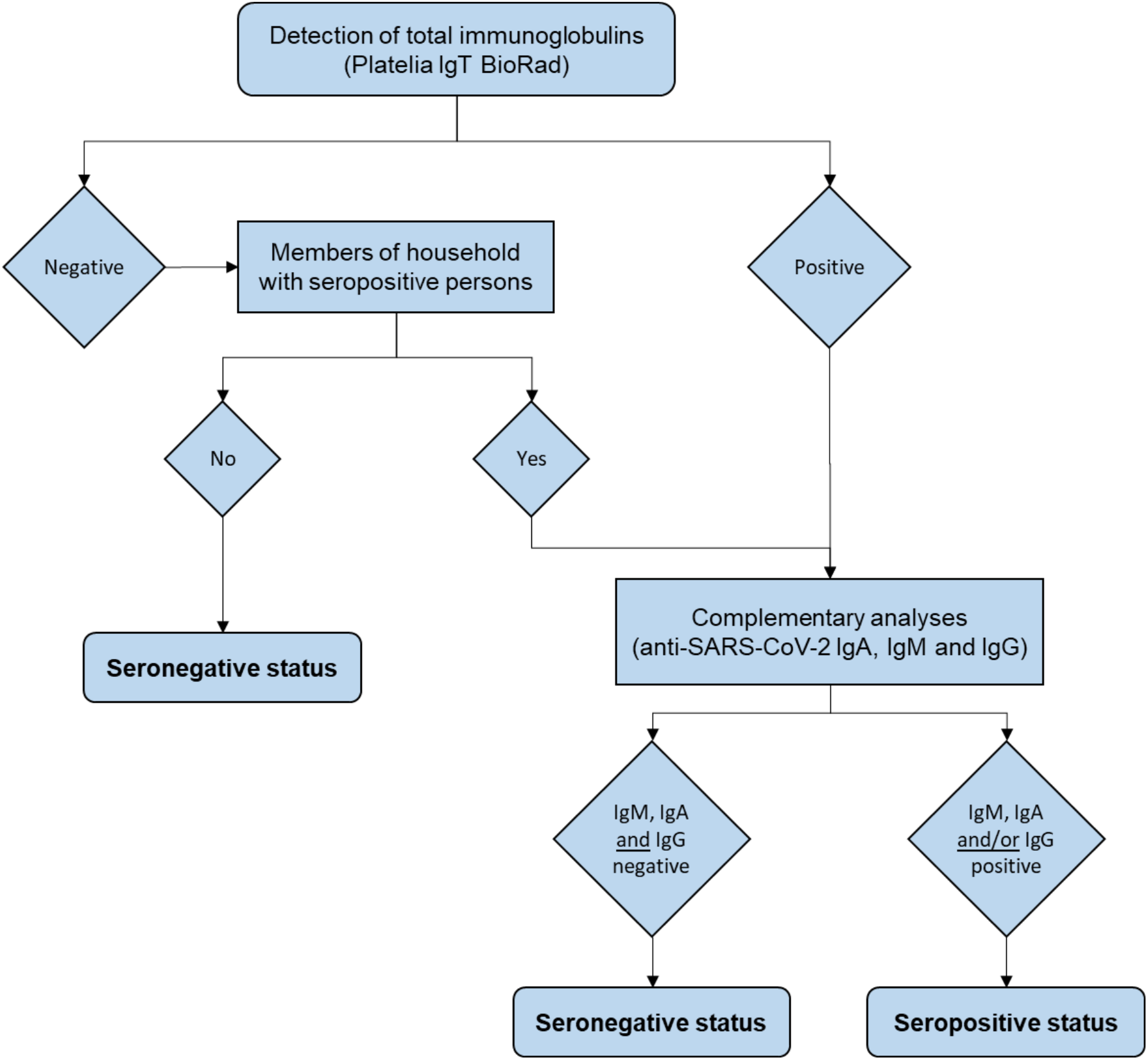
Flow of serology testing in the study.

### Seroneutralization assay

The SARS-CoV-2 strain from a positive respiratory sample (Covi-Lor collection, Nancy University hospital, France) was cultured on Vero CCL-81 cells (provided by L2CM Laboratory, Nancy). Sera positive for anti-SARS-Cov-2 antibodies were diluted from 1/10 to 1/640 and incubated with virus suspension for 2 hr. Cells were inoculated with the final suspension. Each dilution was tested five times in the same experiment and each sample in two independent experiments. The cytopathic effect was read on day +6.

Negative controls were uninfected cells; positive controls were the virus incubated without sera and the virus incubated with SARS-CoV-2–negative sera at a 1/10 ratio.

The samples were classified according to neutralization activity at the 1:40 dilution: neutralization > 50% (NT50).

### Statistical analysis

All statistical analyses involved using R 3.6.0. To calculate the 95% confidence interval for fractions, we used the normal approximation interval except for the Clopper Pearson exact method based on binomial distribution for the SARS-CoV-2–positive sample (too small size). The raw seroprevalence estimate was adjusted for age, sex, and EDI quintile, then standardized to the metropolitan and national population (24). For comparing seroprevalence or characteristics between groups, we used chi-square or Fisher exact test and logistic regression, estimating odds ratios (ORs) and 95% confidence intervals (CIs). We used the R package ClustOfVar (25) to study the clustering of symptom variables and draw dendograms. Intra-household infection spread was tested by a permutation test (26). The principle was to generate, by simulation, the empirical distribution of the number of infected households under the null hypothesis, respecting the number of individual cases and the structure of the households observed in the sample. We used simulation to calculate the relative risk (and 95% CI) of being SARS-CoV-2–positive in a household with a SARS-CoV-2–positive member.

## RESULTS

### Sample description (Figure 2)

We invited 6094 people to participate to enable the inclusion of 2006 participants, aged 5 to 95 years old from 1111 households: 55% were women and 148 under 18-year-old; 469 people came to the visit alone, 938 came as a couple and the others (599) came as a family of 3 to 6 people.

**Figure 2:**
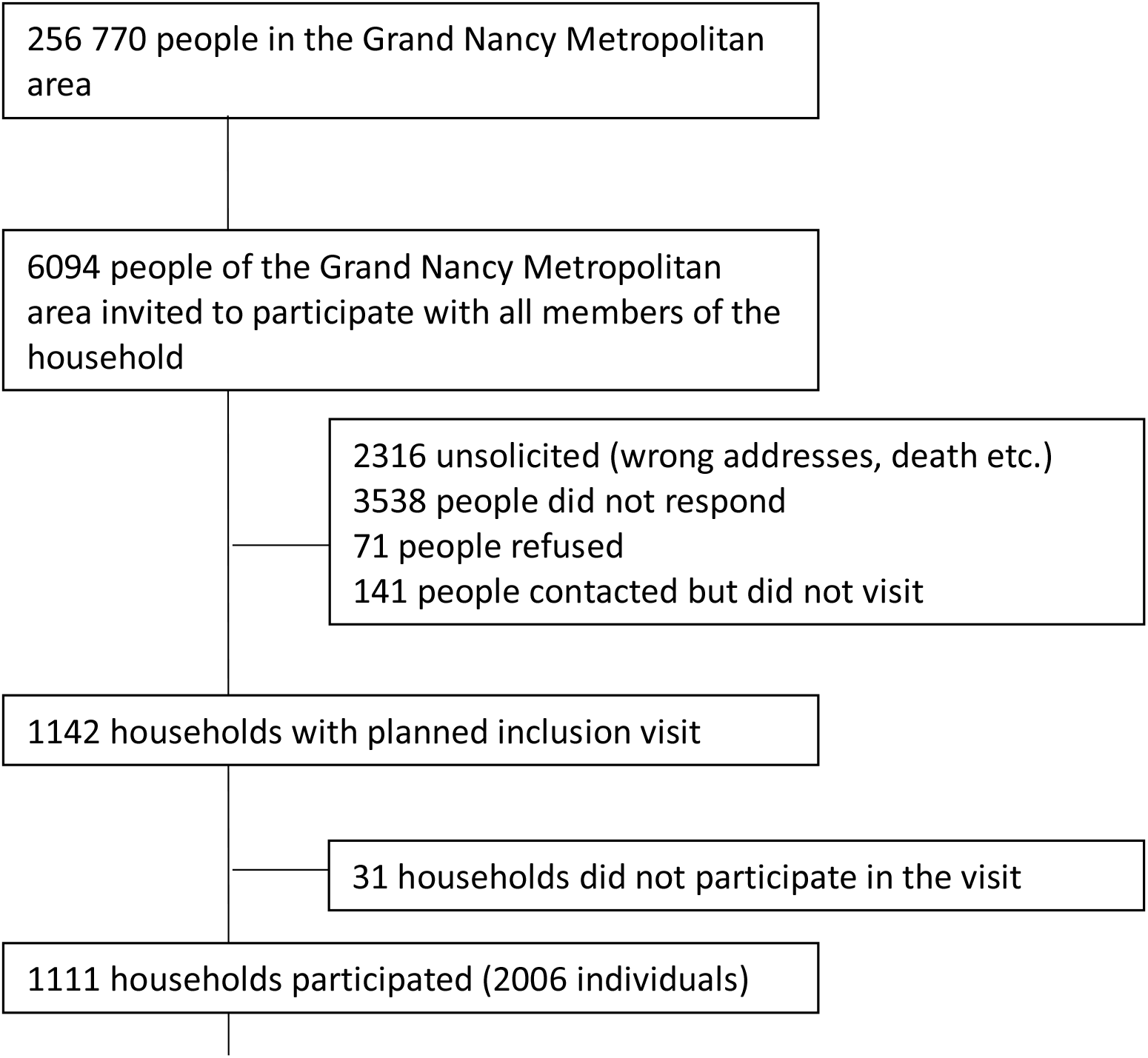
Flow of participants in the study.

The Grand Nancy Metropolitan area comprises 110 IRIS zones; 108 were represented. People in neighbourhoods with a high socio-economic level (measured by the EDI) and high socio-professional category responded better than others. According to the EPICES score, 388 of the 1816 (21%) participants with this score were considered to live in social precariousness.

Social precariousness was also linked to the IRIS EDI quintile: less than 16% in the first three quintiles, up to 23% in the fourth quintile and 40% in the last quintile (p<10^−5^); it also increased with age: 16% in the 5-44 age group, 18% in the 45-64 age group and 28% in those over 65 (p<10^−6^).

Among the 2006 participants, 16% were smokers, 2% used nicotine substitutes, and 29% were former smokers. Moreover, 294 (14.6%) reported at least one comorbidity (among: hypertension, cancer, diabetes, kidney failure, liver problems, immune deficiency, immunosuppressive therapy, severe obesity). The presence of a comorbidity was not related to EDI score but was strongly related to social precariousness: 26% of those in precarious situations had at least one comorbidity as compared with 13% of others (p<10^−9^).

In total, 252 (12.6%) participants thought they were infected with COVID-19 because they experienced symptoms (86%) and/or had been in contact with a sick person (44%). Among contacts with COVID-19, 42% were from work areas, 28% were family and 22% were friends.

### General seroprevalence

According to the results of anti-SARS-CoV-2 IgT detection and complementary analyses performed as described in Figure 1, 43 of the 2006 participants were found to be seropositive. Thus, seroprevalence was 2.1% (95% CI 1.5 to 2.8]). On adjustment for age, sex and EDI quintile, seroprevalence was 2.5 (1.8 to 3.3) standardized for the Grand Nancy Metropolitan area and 2.3 (1.7 to 3.1) standardized for France.

Among the 43 SARS-CoV-2–positive samples, none was positive for anti–SARS-CoV-2 IgM antibody only, 17 (39.5%) were positive for anti–SARS-CoV-2 IgM and IgG antibodies and 26 (60.5%) were positive for anti–SARS-CoV-2 IgG antibody only (Table 1).

**Table 1:** Detection of anti-SARS-CoV-2 total immunoglobulin (IgT) antibodies, determination of Ig isotypes and seroneutralisation activity of seropositive sera

| *n* | IgT | IgM | IgA | IgG | Serological status | Seroneutralization capacity (*n*) |
| --- | --- | --- | --- | --- | --- | --- |
| 1923 | Negative | NR | NR | NR | Seronegative | NE |
| 32 | Negative | Negative | Negative | Negative | Seronegative* | NE |
| 8 | Positive | Negative | Negative | Negative | Seronegative | NE |
| 2 | Negative | Negative | Positive | Positive | Seropositive* | **0** |
| 14 | Positive | Positive | Positive | Positive | Seropositive | **11** |
| 0 | Positive | Positive | Positive | Negative | Seropositive | NE |
| 3 | Positive | Positive | Negative | Positive | Seropositive | **1** |
| 17 | Positive | Negative | Positive | Positive | Seropositive | **15** |
| 0 | Positive | Positive | Negative | Negative | Seropositive | NE |
| 0 | Positive | Negative | Positive | Negative | Seropositive | NE |
| 7 | Positive | Negative | Negative | Positive | Seropositive | **4** |
| 2006 | Total |  |  |  |  |  |

\* Living with seropositive people
NE: not evaluated

### Seroprevalence by age and socioeconomic status (Table 2)

Seroprevalence was highest in the 20-34 age group (4.7% [95% CI 2.3 to 8.4]) and in people from area with lower socioeconomic level (2.7% vs 1% for EDI quintiles 3, 4 and 5 vs 1 and 2, *P=*0.02). We observed little difference in prevalence among people without and with a baccalaureate diploma (1.4% vs 2.6%, *P=*0.10) and social precariousness (1.0% vs 2.5% for EPICES scores >30 vs ≤ 30, *P=*0.09). Social precariousness had no protective effect as seen by the probability of precariousness estimated with adjusted or random-effects models (*P=*0.07 to 0.11) (see Table 2 for one of the models).

**Table 2:** Number of cases, seroprevalence for each risk factor modality. Data are 95% confidence intervals (CIs), odds ratios (ORs) and p values.

| Modalities | Positive<br>/Total | % | % CI | OR | OR CI | P |
| --- | --- | --- | --- | --- | --- | --- |
| **Age** |  |  |  |  |  |  |
| 05-19 | 2/203 | 1.0 | 0.1 - 3.5 | 0.7 | 0.1 - 3.2 | 0.65 |
| 20-34 | 10/215 | 4.7 | 2.3 - 8.4 | 3.4 | 1.2 - 10.9 | **0.03** |
| 35-49 | 5/350 | 1.4 | 0.5 - 3.3 | ref |  |  |
| 50-64 | 16/553 | 2.9 | 1.7 - 4.7 | 2.1 | 0.8 - 6.3 | 0.16 |
| 65-79 | 9/573 | 1.6 | 0.7 - 3.0 | 1.1 | 0.4-3.6 | 0.86 |
| 80+ | 1/112 | 0.9 | 0.0 - 4.9 | 0.7 | 0.0 - 3.9 | 0.67 |
| **Gender** |  |  |  |  |  |  |
| Female | 24/1104 | 2.2 | 1.4 - 3.2 | ref |  |  |
| Male | 19/902 | 2.1 | 1.3 - 3.3 | 1.0 | 0.5 - 1.8 | 0.92 |
| **quintileEDI** |  |  |  |  |  |  |
| 1-2 | 6/615 | 1.0 | 0.4 - 0.2 | ref |  |  |
| 3-4-5 | 37/1391 | 2.7 | 1.9 - 3.7 | 2.8 | 1.3 - 7.3 | **0.02** |
| **Household size** |  |  |  |  |  |  |
| 1 | 9/364 | 2.5 | 1.1 - 4.6 | ref |  |  |
| 2 | 19/882 | 2.2 | 1.3 - 3.3 | 0.9* | 0.4 - 2.0 | 0.74 |
| >=3 | 15/760 | 2.0 | 1.1 - 3.2 | 0.6 | 0.2- 1.5 | 0.24 |
| **Educational level: baccalaureate** |  |  |  |  |  |  |
| Yes | 33/1266 | 2.6 | 1.8 - 3.6 | ref |  |  |
| No | 8/586 | 1.4 | 0.6 - 2.7 | 0.5 | 0.2 - 1.1 | 0.1 |
| missing | 154 |  |  |  |  |  |
| **Smoking status** |  |  |  |  |  |  |
| Non-Smoker | 38/1583 | 2.4 | 0.2 - 3.3 | ref | ref |  |
| Smoker | 4/338 | 1.2 | 0.3 - 3.0 | 0.5 | 0.1 - 1.2 | 0.17 |
| Missing | 85 |  |  |  |  |  |
| **Body Mass Index** |  |  |  |  |  |  |
| <25 | 22/1162 | 1.9 | 1.2 - 2.9 | ref |  |  |
| [25 - 30[ | 15/551 | 2.7 | 1.5 - 4.5 | 1.5 | 0.7 - 2.8 | 0.27 |
| >=30 | 6/284 | 2.1 | 0.8 - 4.5 | 1.1 | 0.4 - 2.6 | 0.81 |
| missing | 9 |  |  |  |  |  |
| **Comorbidity** |  |  |  |  |  |  |
| No | 38/1726 | 2.20 | 1.6 - 3.0 | ref |  |  |
| Yes | 5/280 | 1.78 | 0.6 - 4.1 | 0.8 | 0.3 - 1.9 | 0.65 |
| **Precariousness (Adjusted on Age, Sexe and EDI)** |  |  |  |  |  |  |
| EPICES<=30 | 35/1428 | 2.5 | 1.7 - 3.4 | ref |  |  |
| EPICES>30 | 4/388 | 1.0 | 0.3 - 2.6 | 0.4 | 0.1 - 1.0 | 0.1 |

### Seroprevalence with other factors

Seroprevalence did not differ between smokers and non-smokers (1.2% vs 2.4%) (*P=*0.18) or by sex, household size, weight status or presence of risk factors (Table 2). Of the 252 participants who thought they had COVID-19, 31 (12%) were SARS-CoV-2–positive and 6 (2%) were negative but lived in a household with a SARS-CoV-2–positive person. Furthermore, 72% of SARS-CoV-2–positive individuals thought they had been infected. Among those who did not think they had COVID-19, 12 (0.7%) were SARS-CoV-2–positive and 20 (1.1%) were negative but lived in a household with a SARS-CoV-2–positive person.

For households, 34 (3.1% [95% CI 2.1 to 4.3]) had at least one SARS-CoV-2–positive individual. Household seroprevalence was slightly higher if the household size was ≥ 3 (4.1% vs 2.7%, *P=*0.26). We found intra-household spread (permutation test, p<10^−6^), and probability of infection was multiplied by 30 (95% CI 11 to 78]) with a SARS-CoV-2–positive household member.

### Symptoms

In the overall sample, 25% (95% CI 23 to 27) showed symptoms that would indicate they had COVID-19. This criterion was related to seroprevalence (6.5% vs 0.7% with and without COVID-19 symptoms, p<10^−13^). Nearly half of the individuals (47%) reported experiencing at least one of the 18 collected symptoms (14% one “intense” symptom). Seroprevalence was higher with than without at least one symptom (3.8% vs 0.7%, p<10^−5^) and when at least one of the symptoms was qualified as “intense” (9.4% vs 0.7%, p<10^−17^). For each of the identified symptoms (except irritated eyes and rash), seroprevalence was higher when the symptom was expressed (see Table 3), with anosmia or agueusia the most discriminating symptom (OR 27.8 [95% CI 13.9 to 54.5]).

**Table 3:** Frequency of symptoms by serology status

| Clinical criterion | Seropositive | Seronegative | p-value |
| --- | --- | --- | --- |
| *n* | 43 | 2006 |  |
|  | % | % |  |
| At least one symptom | 83.7 | 47.6 | 3,E-06 |
| At least one intense symptom | 60.5 | 13.1 | 1,E-18 |
| Clinical criteria poss Covid-19* | 74.4 | 23.8 | 3,E-14 |
| Fever | 62.8 | 14.7 | 1,E-17 |
| Cough | 53.5 | 12.1 | 1,E-15 |
| Fatigue | 48.8 | 10.9 | 6,E-11 |
| Shortness of breath | 46.5 | 6.6 | 6,E-23 |
| Aches | 41.9 | 8.2 | 2,E-14 |
| Anosmia/ageusia | 39.5 | 2.3 | 5,E-44 |
| Muscle pain | 37.2 | 10.4 | 3,E-08 |
| Sore throat | 34.9 | 14.7 | 3,E-04 |
| Headaches | 32.6 | 10.1 | 2,E-06 |
| Rhinorrhea | 30.2 | 16.6 | 0.02 |
| Chest pain | 25.6 | 6.3 | 6,E-17 |
| Diarrhea | 23.30 | 8.4 | 0,0006 |
| Abdominal pain | 20.9 | 6.8 | 0,0004 |
| Loss of balance | 14.0 | 4.0 | 0,001 |
| Nausea | 14.0 | 3.8 | 0,0009 |
| Appetite loss | 11.6 | 1.1 | 2,E-09 |
| Skin rashes | 7.0 | 4.9 | 0.52 |
| Irritated eyes | 4.7 | 6.0 | 0.70 |

\*Definition of ECDC

Focusing on the 43 SARS-CoV-2–positive individuals, 7 (16.3% [95% CI 6.8 to 30.7]) experienced no symptoms. The asymptomatic form did not depend on age (*P=*0.4) or sex (*P=*0.9).

Most symptomatic people experienced symptoms in March (Figure 3), which shows a clear effect of confinement on slowing/stopping the spread of the disease.

**Figure 3:**
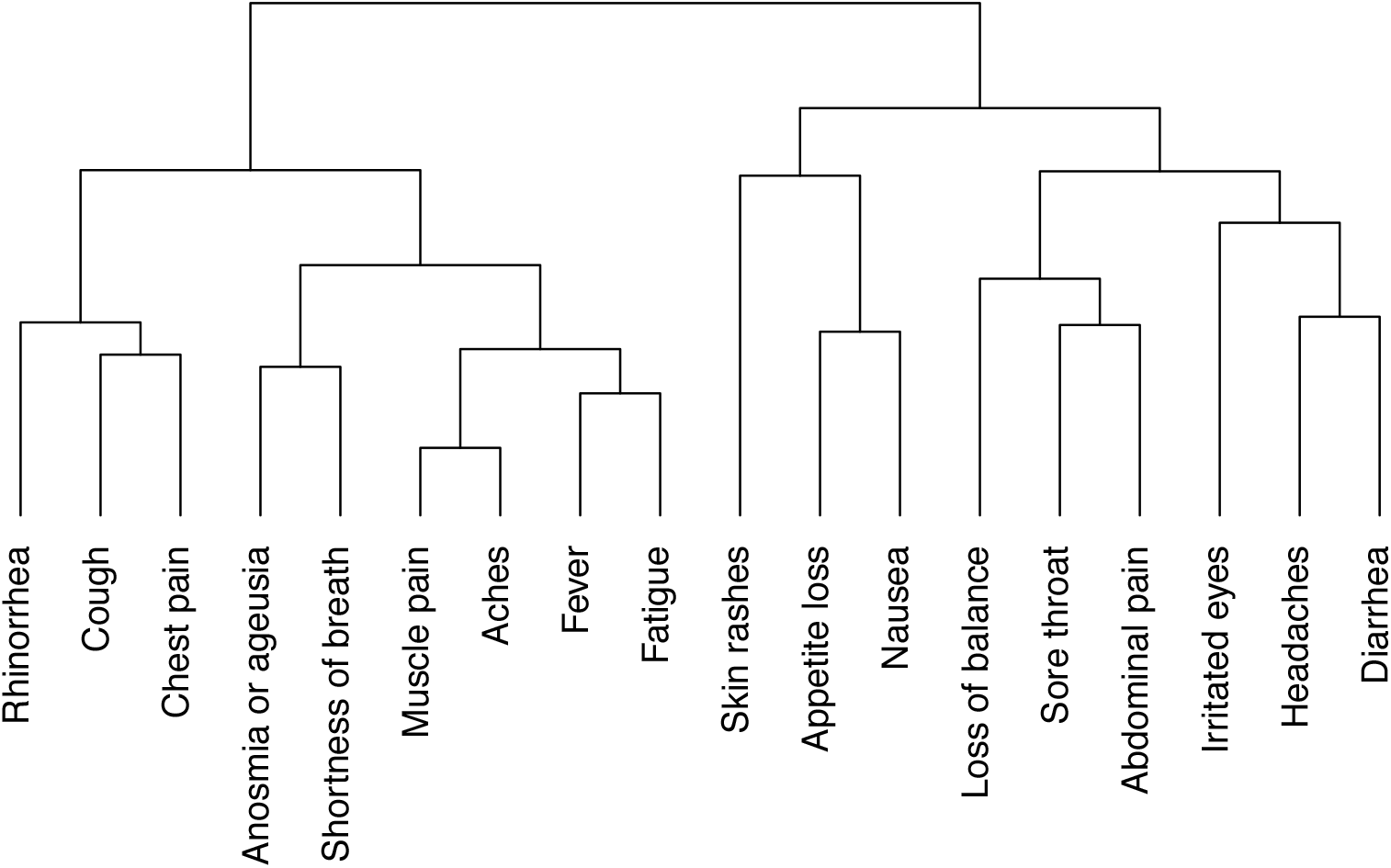
Dendogram of symptoms in seropositive individuals (n=43).

Among those with at least one symptom, many (72%) reported this symptom as “intense”. A cluster analysis of symptoms in SARS-CoV-2–positive individuals grouped anosmia or agueusia with influenza-like illness (Figure 4). Anosmia or agueusia was strongly associated with shortness of breath (*P=*0.0002) and fever (*P=*0.0008) but almost never occurred without fever (1/17).

**Figure 4:**
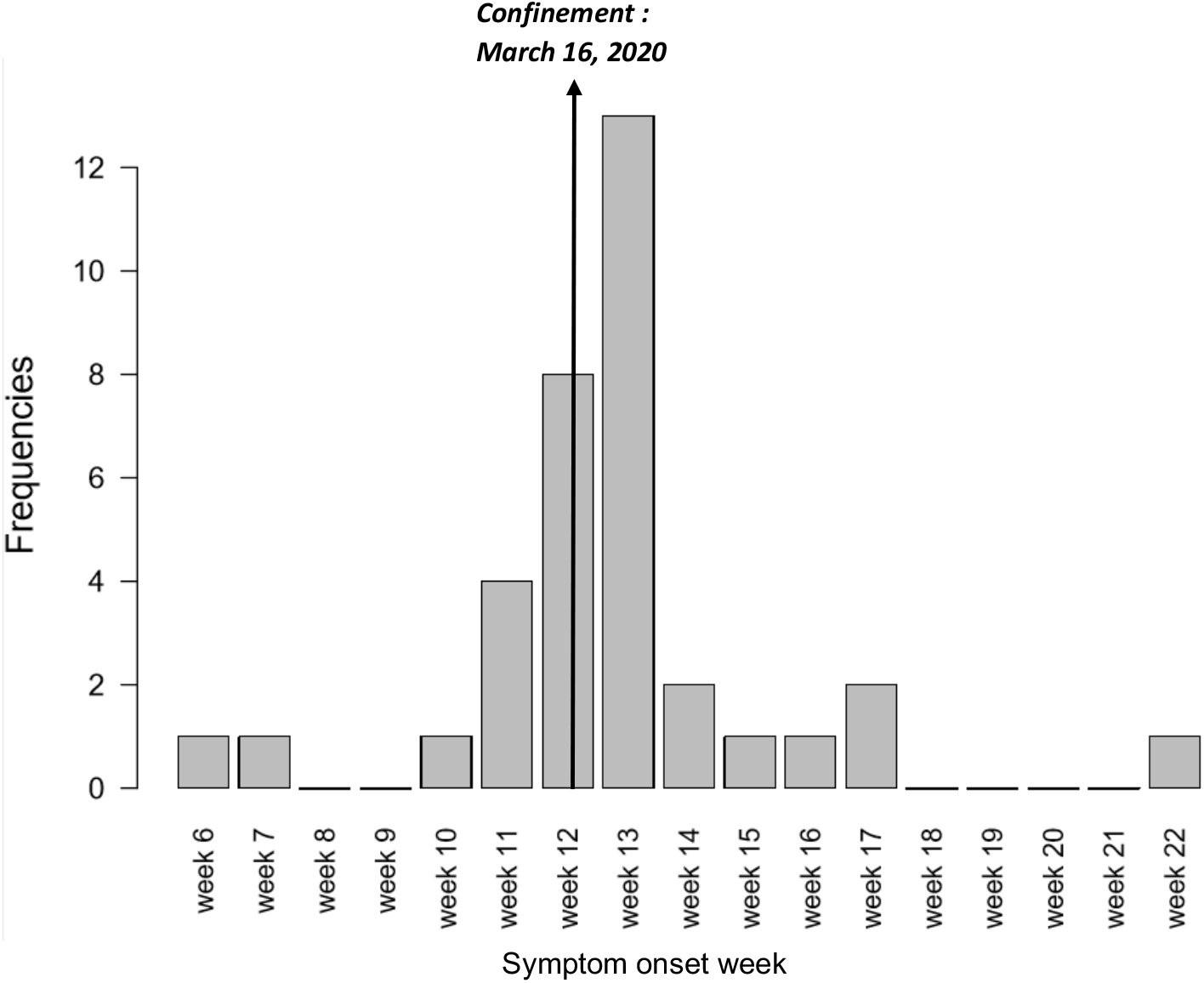
Date of onset of symptoms in seropositive symptomatic individuals (n=36).

### Seroneutralization assay

For 31/43 (72% [95% CI 56 to 85]) SARS-CoV-2–positive individuals, antibody detection was associated with neutralization activity (NT50 ≥ 40) (Table 1). Twelve seropositive samples presented no or weak neutralization capacity. Among 17 serum samples with recent seroconversion (IgM+IgG positive), 12 (71%) presented seroneutralisation activity, as did 19/26 (73%) with older seroconversion (IgG only) (NT50 ≥ 40). Seroconversion did not depend on age (*P=*0.8), sex (*P=*0.6), or time between symptoms and data collection. Seroneutralization did not depend on the presence of symptoms (*P=*0.9). The NT50 was increased with strong symptoms (*P=*0.24).

## DISCUSSION

In July 2020, the low seroprevalence of SARS-CoV-2 (i.e., 2.1% SARS-CoV-2–positive cases among the 2006 included individuals) underlined how the Grand Nancy Metropolitan area population remained immuno-naïve and susceptible to the second epidemic wave that occurred during autumn/winter 2020/2021 in France. One in six SARS-CoV-2–positive individuals remained asymptomatic.

Seropositivity was associated with the ecological marker social deprivation (EDI) but not when assessed with social precariousness at the individual level (EPICES), which may indicate some protective effect. Such discrepancy might be related to the social isolation of deprived individuals during the lockdown, independent of residence, potentially resulting in less exposure for many reasons (unemployment, fear of meeting people).

Within the *Coronaviridae* family, SARS-CoV-2 is phylogenetically close to SARS-CoV and to a lesser extent MERS-CoV (27). Cross-reactivity with seasonal coronaviruses has to be discussed. Even if recently, pre-existing HCoV-NL63 antibody response cross-reacting with some SARS-CoV-2 antigens was detected in both pre- and mid-pandemic infected individuals (28), no cross-reactivity of serology methods with the seasonal coronavirus was reported (29) and no neutralizing activity of SARS-CoV-2 in pre-pandemic sera was observed (30). All positive serum samples were positive for antibodies against the RBD of S protein (Biosynex test). IgM and IgG antibodies directed against the RBD are assumed to decrease in titers during the 6 months post-infection. Since the study took place less than 6 months after the first epidemic peak, the seroprevalence is representative of the exposure of the population to SARS-CoV-2 during the first wave. The question of the remaining humoral immunity is currently unsolved, but even though the IgG titers and neutralizing activity decreases, the number of RBD-specific memory B cells was unmodified at 6 months, which can contribute to the immune response to a secondary infection (31). Moreover, the presence of high IgG and IgM antibodies to the spike S1 C-terminal domain in recovered patients might be associated with efficient immune protection in COVID-19 patients (32).

A recent work reported that standard commercially available SARS-CoV-2 IgG results could be a useful surrogate for neutralizing antibody testing (32). However, in the present study, antibody neutralizing titers were determined *in vitro* by using a native SARS-CoV-2 strain to be closer to the physiological infection of cells by SARS-CoV-2 in northeast France (33). Twelve of 43 SARS-CoV-2–positive individuals presenting no or weak neutralizing titers agrees with a study of COVID-19 recovered patients (33).

This study has strengths. First, in this relatively small geographical area, we were able to stratify sampling based on homogenous population zones (IRIS zones) and tag the level of social deprivation by using the corresponding ecological EDI index, to better represent the target population in terms of this variable that has been considered an important risk factor for COVID-19 (8). Second, seroprevalent cases were carefully identified by using several methods for confirming seropositivity (figure 1). Third, seroneutralization capacity was investigated in duplicate, in line with recommended standard practices, instead of derived from IgG results (33,34)

The study has some limitations. First, the sampling electoral database, chosen for its immediate availability in such a small and delimited area, did not completely cover the adult population because some people left the area (among the 6094 people invited, 38% were not really solicited [wrong addresses, death etc.]), newcomers to the area did not yet register (not mandatory), and non-European citizens were not eligible, which creates some representativeness bias (35). Second, the estimated response rate was relatively low in a period immediately following the lockdown, with many people already gone away for July summer holidays. Third, all data were self-reported, which may lead to some measurement (declaration) bias. Moreover, even if the number of individuals was sufficient to satisfy the main objective, the study lacked statistical power for the study of risk factors.

Finally, with novel SARS-CoV-2 variants emerging all over the world (36), the neutralizing activity of positive sera against novel SARS-CoV-2 variants could be evaluated.

In conclusion, IgT assays are key tools to monitor the circulation of SARS-CoV-2 and the impact of public health guidelines (37). In this population of low anti-SARS-CoV-2 seroprevalence, a beneficial effect of the lockdown can be assumed with frequent SARS-CoV-2 seroneutralization among IgT-positive patients. IgT seroprevalence was higher for young adults and was associated with intra-family SARS-CoV-2 transmission.

## Data Availability

The data for this article will be shared on reasonable request to the corresponding author.

## DATA AVAILABILITY STATEMENT

The data for this article will be shared on reasonable request to the corresponding author.

## ACKNOWLEDGMENTS

The authors warmly acknowledge the inhabitants of the Grand Nancy metropolitan who participated in COVAL Nancy Study. COVAL is funded by the Grand Nancy metropolitan. The sponsor was CHRU de Nancy (Direction de la Recherche Clinique et de l’Innovation).

Many people worked together enthusiastically to make the COVAL Nancy study a success. **For Université de Lorraine**, we would like to thank Nicolas THORR, Julia Budzinski and Didier GEMMERLÉ (Institut Élie Cartan de Lorraine) for participating in the draw and statistical analysis.

Damien GARAUD for his open source code of Iris geolocation.

**For CHRU de Nancy**, we would like to express our gratitude to:

- the Département Méthodologie, Promotion, Investigation, Mehdi SIAGHY, Nathalie THILLY, Marjorie STARCK and Louise GEND for their support.
- the Centre d’Investigation Clinique Epidémiologie Clinique members for their contribution to organisation, blood samples and data collection, data entry and management, activity reporting, and logistic with Sylvie KLEIN Marie SPONGA, Jean-Marc VIRION, Laurence EMPORTE, Sandrine GRANDCLERE, Emilie JACQUOT, Sylvie RONCHETTI Agathe BOCHNAKIAN, Heloise BRIANCON, Alfousseyni COLY, Marie Rita LETOURNEUR, Sandrine GERSET, Nathalie DUMONT, Nathalie PIERREZ, Nina THIAVILLE, Sandrine TYRODE, Véronique VOGEL, Christelle DUJON, Nicole KOEBEL, Philippe MELCHIOR and Samia MAHMOUDI.
- the Centre d’Investigation Clinique Pluridisciplinaire members for their contribution to organisation and realisation of blood samples: Patrick ROSSIGNOL, Nicolas GIRERD, Edith DAUCHY, Helene COYARD, Evelyne MICOR, Agnes PEZZI, Lydie POINSIGNON and Jonathan UDOT.
- the Unité d’Investigation Clinique for their contribution to organisation and realisation of blood samples: Nathalie THILLY, Valerie BOUAZIZ and Marie Alexandra PAQUOT et Nelly FRANCOIS
- the Hôpital d’Enfant, Cyril SCHWEITZER, Valerie RATAJCZAK, Herve LEROY the and the pediatry nurses for the realisation of child blood samples
- the technical and management teams of the Laboratories for the realization of SARS-CoV-2 serology and samples management, Mihayl VARBANOV and Stéphanie PHILIPPOT of the L2CM Laboratory (Nancy) for the cell model of culture of SARS-CoV-2 and Jonathan MAYER (medical student) for his contribution to the performance of seroneutralization analyzes.
- the Direction des Soins in particularly Mireille GAUDRON, Sandrine HAYO VILLENEUVE, Sandrine JORAY et Julie THOUVENIN-GALANTI
- the Centre de Ressources Biologiques (CRB) Lorrain of Nancy BB-0033-00035 for managing patient samples : Catherine MALAPLATE and Sandra LOMAZZI.
- the Communication members Laurence VERGER and David KOZON
- the Direction des Services Informatiques in particularly Lionel SCHWEITZER

